# Effect of vaccination rates on the prevalence and mortality of COVID-19

**DOI:** 10.1101/2022.03.31.22273274

**Authors:** Jacob Westerhout, Hamid Khataee, Zoltan Neufeld

## Abstract

By looking at trends in global epidemic data, we evaluate the effectiveness of vaccines on the incidence and mortality from the delta variant of COVID-19. By comparing countries of varying vaccination levels, we find that more vaccinated countries have lower deaths while not having lower cases. This cannot be explained by testing rates or restrictions, but can be partly explained by the most susceptible countries also being the highest vaccinated countries. We also find that during the period when many countries have high vaccination rates, cases and deaths are both increasing in time. This seems to be caused by the waning of the protection vaccines grant against infection.

## Introduction

During the first year of the COVID-19 pandemic, non-pharmaceutical interventions that limit social interactions and mobility (such as strict lockdowns) were implemented to control the spread of the disease [1]. By late 2020, a number of COVID-19 vaccines showed efficacy in preventing COVID-19 in phase 3 clinical trials [2] and mass vaccinations began across the world.

Vaccines have been shown to be effective against asymptomatic and symptomatic infection, severe infection, hospitalisation, and death [2–8], but the containment of the disease is challenged by a number of factors.

One potential cause of outbreaks is the emergence of new variants [9]. Vaccines tend to be less effective at protecting against infection from certain variants, but are still effective against severe outcomes [10, 11].

A second potential cause of outbreaks in vaccinated populations is the waning of the immunity granted by vaccinations. It has been shown that the protection against infection waned a few months after receiving the second dose, but the protection against hospitalization and death persists [3, 4, 12–14].

To investigate the relationships between infections and vaccination rates at the global-level, correlation analyses have been performed on worldwide time series data. For example, data from 91 countries showed no correlation between the vaccination level and daily new cases [15]. In a similar analysis, increases in daily new cases were found to be unrelated to levels of vaccination, based on data from in 68 countries and 2947 counties in the United States [16]. These findings reveal a need for a comprehensive analysis of global data to provide a worldwide representation of the effects of health measures on suppressing the COVID-19 disease.

In order to address some of the potential limitations of these previous studies (see [17–19]) we analyse a larger data set covering 190 countries over multiple time-windows within a timeframe of several months duration. Our data analysis also considers the broader context including the number of daily tests, level of restrictions, human development index, and the number of COVID-19 vaccine doses received per person as covariates of daily cases and deaths per capita. To limit the variability caused by different variants we focus on the spread dynamics of COVID-19 in vaccinated populations within the time-period when the delta variant was dominant. For example we do not include the omicron variant for which there is insufficient data on severe disease outcomes [20] and with very high daily cases the data on the spread of infections is less reliable due to limited testing capacity. We also mainly focus on the period when vaccination rates are sufficiently high as low vaccination rates are unlikely to significantly alter the pandemic.

## Data and Analysis

The data were collected from multiple COVID-19 data resources for 190 countries from 1-March-2021 to 1-December-2021.

We sourced the number of daily tests from Our World In Data [21]. Johns Hopkins University repositories were used to extract the number of daily cases and deaths [22] and the number of vaccines distributed [23]. The Oxford COVID-19 Government Response Tracker data [24] was used to source the stringency index, a composite measure of the level of restrictions scaled to values from 0 (no restriction) to 100 (strictest). Data form United Nations Development Programme [25] was used to extract human development index (HDI, composite metric of life expectancy, education, and living standard [26]), where HDI *>* 0.8 and HDI *<* 0.55 represent high and low development measures respectively [27]. The data for the population of countries were sourced from the World Bank Data [28] for analysis per capita.

To capture important trends in data, instead of looking at trends at a specific time we look at trends within a three month period. This period is small enough to still see the dynamics of the pandemic but large enough to minimise the effects of small outbreaks. To show these trends we use a moving average line. The value of the line at a point *x* is calculated as the average of all data lying within an interval centred around *x*. The length of the interval depends on the data being plotted and is chosen arbitrarily so the line does not seem noisy but also not over-smoothed. While the average of the data is plotted for clarity, the moving average line is calculated using all data points.

## Results

We find that daily cases per capita, calculated as an average across a three months time window, varies considerably for countries of very similar vaccination levels, indicating that the vaccination rate is not a dominant predictor of cases. We also find that above moderate vaccination levels there is no clear relation increase or decrease in the average number of vaccine doses received per person and the daily cases per million; see Figure 1(a-c). Above a certain vaccination level there is a clear trend that countries with higher average vaccine doses received per person have lower daily deaths per million; see Figure 1(b-d). This then indicates that deaths per case also tends to decrease with vaccination; see Figure S4. The noise in deaths also reduces as vaccination increases indicating that vaccinations are a dominant predictor of deaths at high vaccination levels.

**Figure 1:**
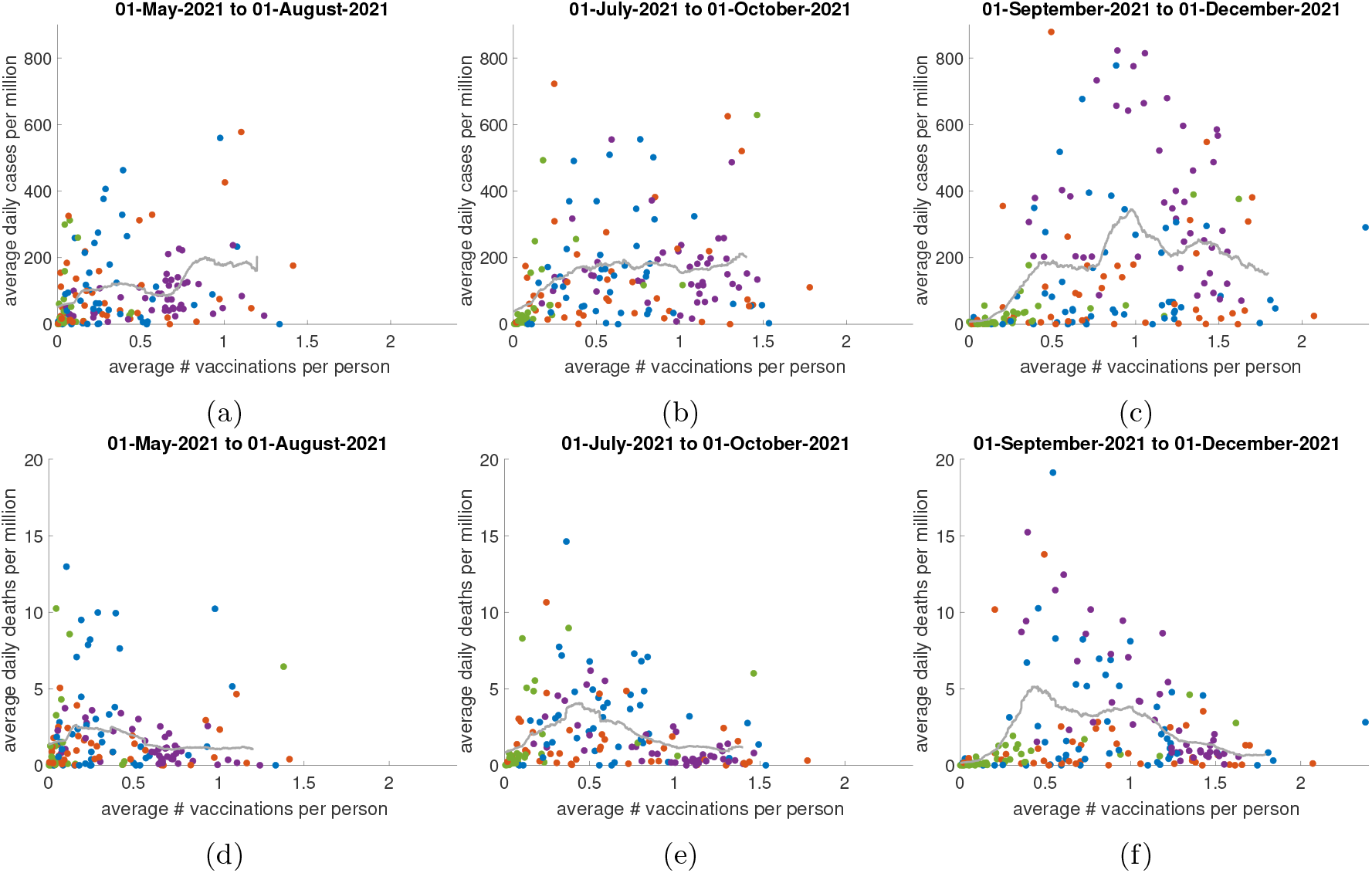
Average daily cases (a-c) and deaths (d-f) versus average number of vaccine doses received per person in 190 countries (data points). Values are averaged over the period between the dates titled. Colour scheme: red (Asian countries), purple (European countries), green (African countries), and blue (all other countries). Grey line represents the moving average line for intervals of length 0.2.

There is a sharp increase in both cases and deaths at low vaccination level. The vaccination level where this sharp increase stops is not constant in time, indicating that vaccines are not the cause. There is an overall increase in cases in time despite the increasing vaccination levels. The average daily cases per million in Figures 1(a-c) respectively are 93.4, 125.5, and 155.1. There is also an overall increase in deaths in time with the average deaths in Figures 1(d-f) being 1.5, 1.7, and 2.1. The increase in cases seems to be roughly independent of the vaccination level while the increase in deaths is mostly concentrated in the lower vaccinated countries. This is consistent with the evidence that vaccine’s protection against infection wanes in time but their protection against death does not.

In order to investigate the source of these observations described above we looked at the relationship to restrictions. We found no significant relationship between the stringency index (the measure of restrictions) and vaccination rates, see Figure 2(a-c)), and hence restrictions are not the cause of any of the trends given in Figure 1.

**Figure 2:**
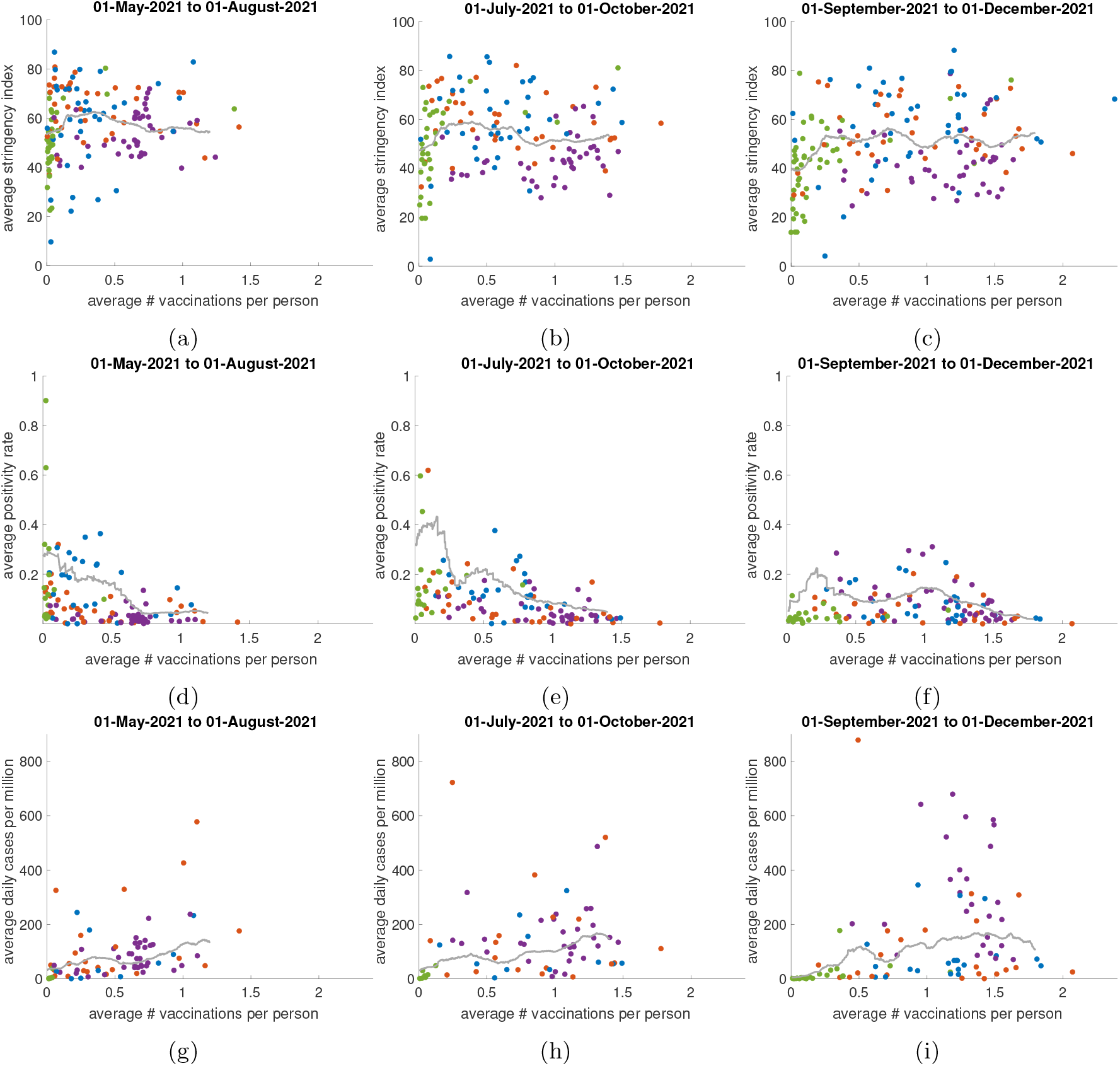
Average stringency index (level of restrictions) (a-c), average positivity rate (ratio of cases to tests) (d-f) and daily cases per million for countries with a positivity rate (ratio of cases to tests) of less than 0.1 (indicating adequate testing) (g-i) versus the number of vaccine doses received per person for all countries with data (each data point). Values are averaged over the period between the dates titled. Colour scheme: red (Asian countries), purple (European countries), green (African countries), and blue (all other countries). Grey line represents the moving average line for intervals of length 0.2.

Restrictions on average are decreasing in time with he average stringency index in Figures 2(a-c) respectively being 52.8, 50.4, and 47.3. This could partially explain the lack of a decrease over time in average cases between Figure 1(a-c) but cannot be the driving cause as the decrease is minor and also cannot explain the lack of decrease of cases with vaccination rates. Comparing testing rates across different countries there is a clear trend of more vaccinated countries doing more tests (see Figure S1), but this itself does not mean that the trends in Figures 1(a-c) are caused by testing rates. It is possible that the countries that test more do so because they indeed have more cases. This can be determined using the positivity rate: the ratio of detected cases to tests administered. A high positivity rate indicates that the true number of cases is much higher than the reported number [29].

It is the case that generally lower vaccinated countries have a higher positivity rate and hence more unreported cases; see Figure 2(d-f). This then seems to explain the lack of a decreasing trend in cases at high vaccination levels. However it is impossible to tell precisely how much higher the true case numbers are just from the positivity rate. When only considering countries with positivity rate below a certain threshold the trends are still similar to those given in Figure 1(a-c); see Figures 2(g-i) and S2.

A significant missing covariate in Figure 1 is the wealth of countries. Figures 3(a) and S3(a) show that the wealthier a country is, the higher average vaccinations per person. This is partly caused by wealthier countries purchasing more vaccines [30].

**Figure 3:**
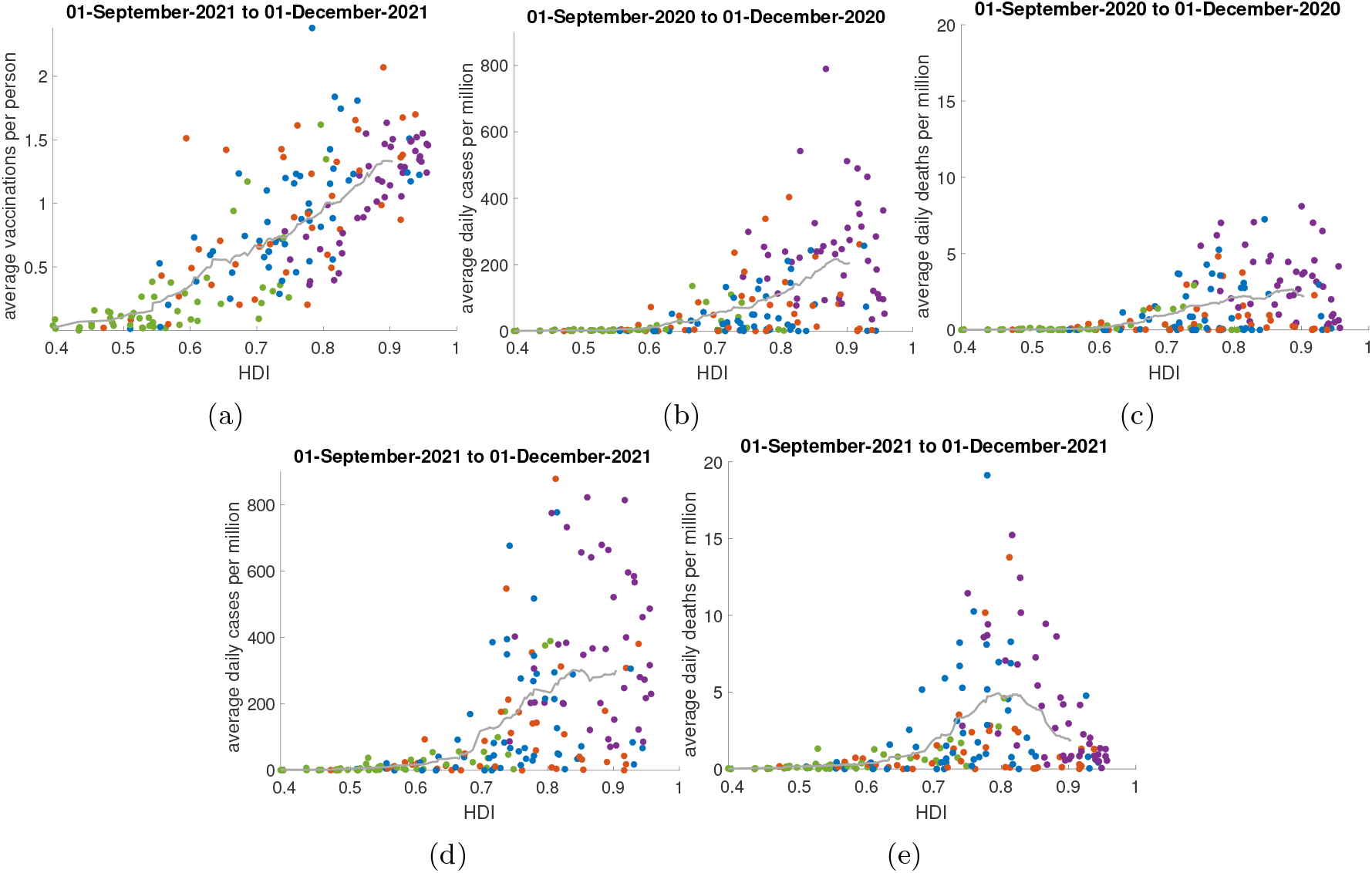
Human development index (HDI, the net wealth per capita) vs the average number of vaccines doses received per person (a), daily cases per capita (b-d), and daily deaths per capita (c-e). Values are averaged over the period between the dates titled. Colour scheme: red (Asian countries), purple (European countries), green (African countries), and blue (all other countries). Grey line represents the moving average line for intervals of length 0.1.

Figures 3(b,c) and S3(b,c) show that pre-vaccinations, the wealthier a country is the higher cases and deaths on average. Even before before mass testing a similar result was hypothesised using on-the-ground realities. [31]. This is unexpected as this is the first time in post-war history that poor countries are not the most affected by a pandemic [32].

The reason for this is unknown, although there are a few theories. Some possible significant covariates are the average yearly number of people arriving per capita [33, 34], median age [34–36], or air pollution [35, 37–41], but none of these appear to provide a convincing explanation for the correlation between wealth and cases or deaths.

More recently it is still the case that wealthier countries have more cases; see 3(d). However, recently above around 0.8 HDI wealthier countries have a decreasing trend for deaths; see 3(e).

This then means that the portion of Figure 1 with increasing trend in cases and deaths can be explained by the more vaccinated countries being more susceptible to the virus. The downwards trend in deaths in Figures 1(d-f) cannot be explained by wealth and therefore is likely caused by vaccination.

## Conclusion

We find significant evidence that vaccines are effective at reducing deaths from COVID-19, but do not find significant evidence that vaccines are effective at reducing the prevalence of infections in the population. This seems to contradict the clinical evidence that vaccines are effective in reducing the chance of infection at individual level. A likely explanation for this contradiction is the waning of the protection vaccines grant against infection.

It seems that vaccines cannot end the pandemic, but they can likely reduce the deaths caused by the pandemic to a very low level of mortality. Although we didn’t analyse data on hospitalisation and severe disease, as such data is not widely available, it is also likely that vaccines could significantly reduce other severe outcomes from the pandemic.

Even if everyone in the world was vaccinated, it seems as though outbreaks would still happen. This is likely even more so the case with the Omicron variant with vaccines being much less effective at protecting from infection [3].

There are implications of our analysis regarding regulations introduced in relation to vaccination against COVID-19. Introducing compulsory vaccination requirements for the overall population (especially for those more vulnerable to the disease) could be justified on the basis that this can reduce the impact on the health care system during an outbreak. However, our results do not support the justification of selective measures restricting the access of unvaccinated individuals to certain venues, universities, travel and workplaces in education, healthcare etc. as this is unlikely to reduce the overall prevalence of the virus within the population.

## Data Availability

Data are sourced from publicly available databases cited in the manuscript.

## Supplementary Material

**Figure S1:**
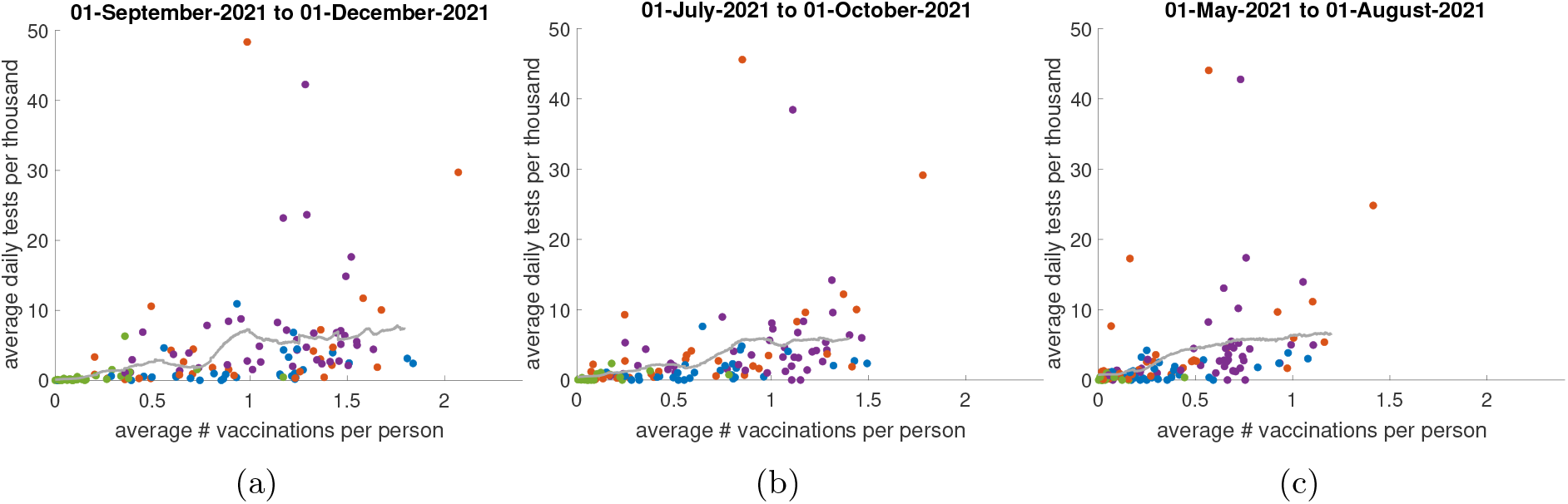
Average daily tests per thousand population versus average number of vaccine doses received per person (data points). Values are averaged over the period between the dates titled. Colour scheme: red (Asian countries), purple (European countries), green (African countries), and blue (all other countries). Grey line represents the moving average line for intervals of length 0.2.

**Figure S2:**
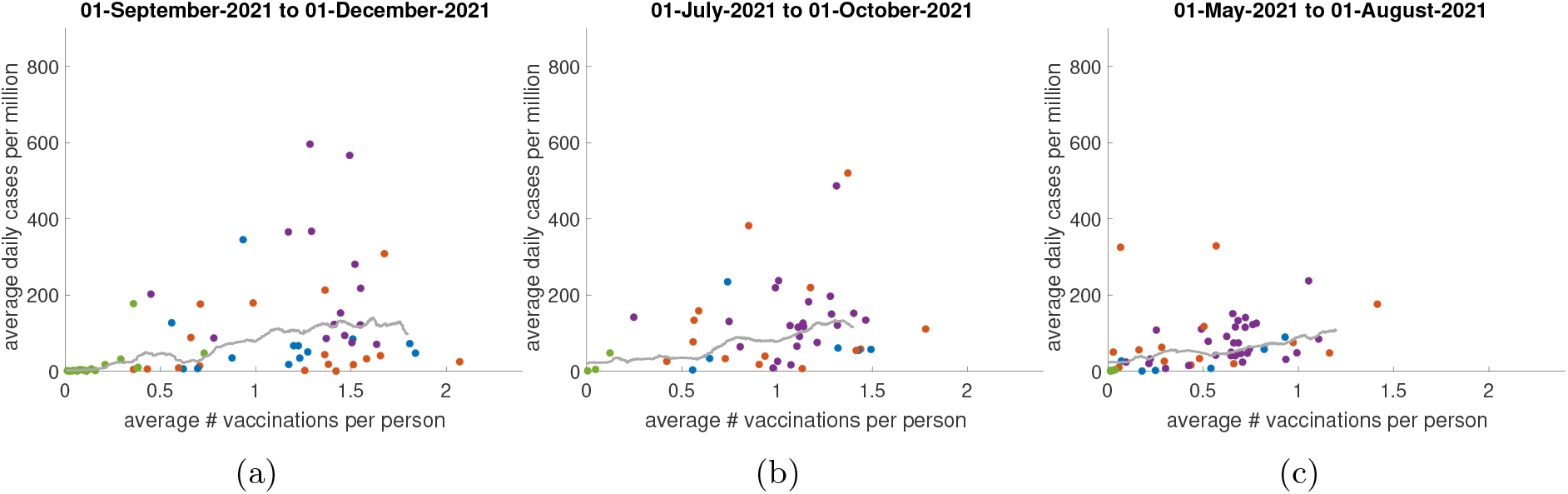
Average daily cases for countries with a positivity rate (ratio of cases to tests) of less than 0.05 (indicating adequate testing) versus average number of vaccine doses received per person in 190 countries (data points). Values are averaged over the period between the dates titled. Colour scheme: red (Asian countries), purple (European countries), green (African countries), and blue (all other countries). Grey line represents the moving average line for intervals of length 0.2.

**Figure S3:**
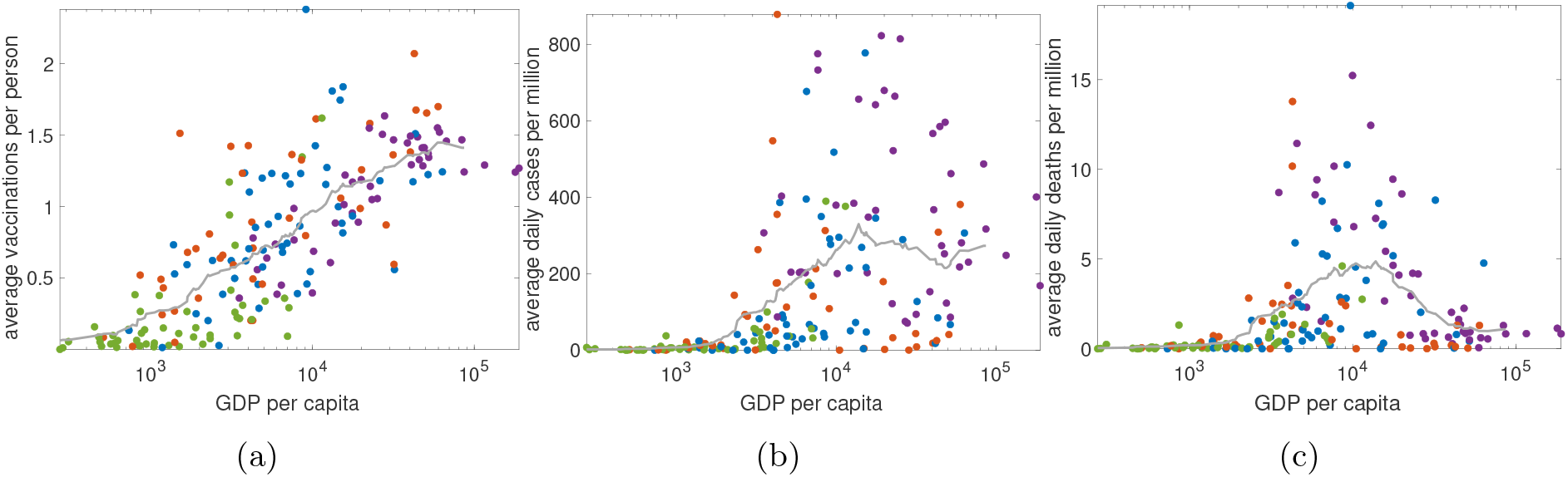
Gross domestic product per capita (GDP, the net wealth per capita) vs the average number of vaccines doses received per person, daily deaths per capita, and daily cases per capita (a-c) respectively. Values in (a) correspond to 1-September-2021 to 1-December-2021 and values in (b,c) correspond to 1-September-2020 to 1-December-2020. Colour scheme: red (Asian countries), purple (European countries), green (African countries), and blue (all other countries). Grey line represents the moving average line for intervals of length 1.2.

**Figure S4:**
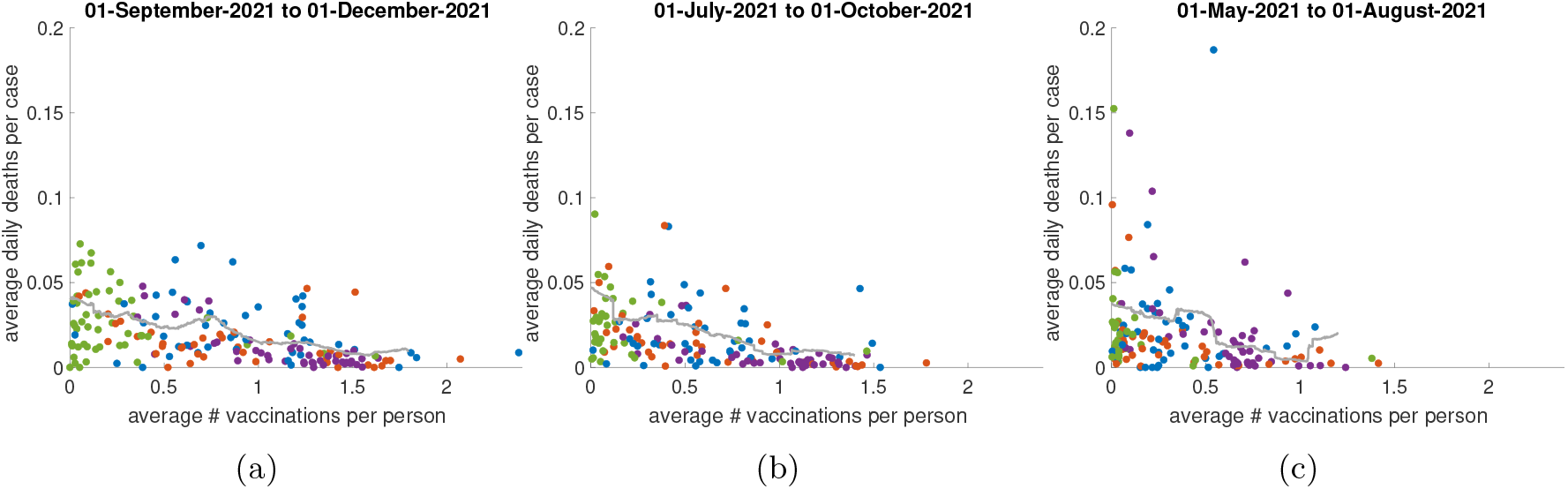
Average daily deaths per case versus average number of vaccine doses received per person (data points). Values are averaged over the period between the dates titled. Colour scheme: red (Asian countries), purple (European countries), green (African countries), and blue (all other countries). Grey line represents the moving average line for intervals of length 0.2.

